# Secondary transmission of COVID-19 in preschool and school settings after their reopening in northern Italy: a population-based study

**DOI:** 10.1101/2020.11.17.20229583

**Authors:** Elisabetta Larosa, Olivera Djuric, Mariateresa Cassinadri, Silvia Cilloni, Eufemia Bisaccia, Massimo Vicentini, Francesco Venturelli, Paolo Giorgi Rossi, Patrizio Pezzotti, Emanuela Bedeschi, the Reggio Emilia Covid-19 Working Group

## Abstract

**Background:** School closures was one of the main measures undertaken to reduce the number of social contacts during the first wave of the severe acute respiratory syndrome coronavirus 2 (SARS-CoV-2) pandemic. We aimed to describe the data on secondary transmission of SARS-CoV-2 among students and teachers/personnel after the reopening of preschools and schools in Reggio Emilia, Italy.

**Methods:** This prospective population-based study included all consecutive cases leading to an investigation in 41 classes of 36 educational institutions (8 infant-toddler centres and preschools, 10 primary and 18 secondary schools) in the period September 1 – October 15, 2020, in Reggio Emilia province, Italy. We report the characteristics of the school, of the index case, including the possible source of infection, the number of contacts (students and teachers/personnel) that were identified and tested and the characteristics of secondary cases.

**Results:** In the study period, 994 students and 204 teachers were tested during related investigations due to notification of 43 primary cases (38 among students and 5 among teachers). Of these, 10 students and two teachers created 39 secondary cases, resulting in an attack rate of 3.9%. There were no secondary cases among teachers/stuff. Secondary transmission occurred in one primary school and 8 secondary schools. Except for two students and one teacher, the possible source of infection for all index cases was identified as they had all had previous contact with a positive case; the majority of secondary cases did not report any previous close contact with a positive case. The clusters ranged from one to 22 secondary cases.

**Conclusions:** Transmission at school occurred in a non-negligible number of cases, particularly in secondary schools. Prompt testing and isolation of classmates could probably reduce the risk of transmission in school settings.

## Background

School attendance is the largest organized source of social contacts in modern society. Therefore, limiting attendance or closing schools is the single action that can immediately reduce the largest number of social contacts. During the COVID-19 pandemic, all efforts to reduce social contacts have been made, and in most countries, these measures have targeted schools as well. These measures have shown to have several consequences on the psychosocial wellbeing of children and adolescents (1-5) and, if prolonged, may have a serious impact on the effectiveness of education in this new generation.

It is therefore important to understand the role of transmission occurring in schools in the spread of the SARS-CoV-2 virus. This task is made more complex because the vast majority of children and adolescents are asymptomatic: in the absence of screening, most infections are not diagnosed and they are rarely identified as index cases in clusters outside of school.

One recent Italian study reported the number of cases occurring in students attending schools (6), suggesting a low incidence among students. However, this figure does not measure the risk of transmission at school since we do not know where those infections were acquired; this figure is mostly determined by the circulation of the virus in that particular moment in the community. A Korean study showed very low transmission rates in kindergartens (7). Thus, the role of schools in virus transmission could be limited (8, 9), meaning that control measures could be oriented elsewhere (10).

Nevertheless, it is important to collect this information in different countries since the risk of transmission likely depends on many characteristics, such as the social distancing measures adopted in that country, the infrastructural and climatic contexts and the behaviours of children, which are influenced by age and cultural factors.

In this short communication we present the results of 41 investigations conducted after the identification of SARS-CoV-2 infection.

## Methods

### Setting

Reggio Emilia province, northern Italy, has 530,000 inhabitants. The cumulative incidence of cases during the first wave of the pandemic (March – April 2020) was about 0.9%. There are approximately 31000 children attending infant-toddler centres (ages 0-3 years), preschools (ages 3-5), primary schools (ages 6-10), middle schools (ages 11-13) and high schools (ages 14-19). Infant-toddler centres and preschools reopened on September 1, and some special courses in high schools were held, but the official reopening of all schools was on September 15, after three months of very low COVID-19 incidence (figure). In October, the province began experiencing a second wave of the pandemic, with cumulative incidence reaching 1.8%. While the first wave was characterized by very high fatality rate (up to 18% in the first month) (11) and testing limited only to symptomatic individuals, who often presented with severe symptoms, during the endemic period and the beginning of the second wave, most cases have been asymptomatic and identified through active contact tracing. The case fatality rate has dropped to <2%.

**Figure.**
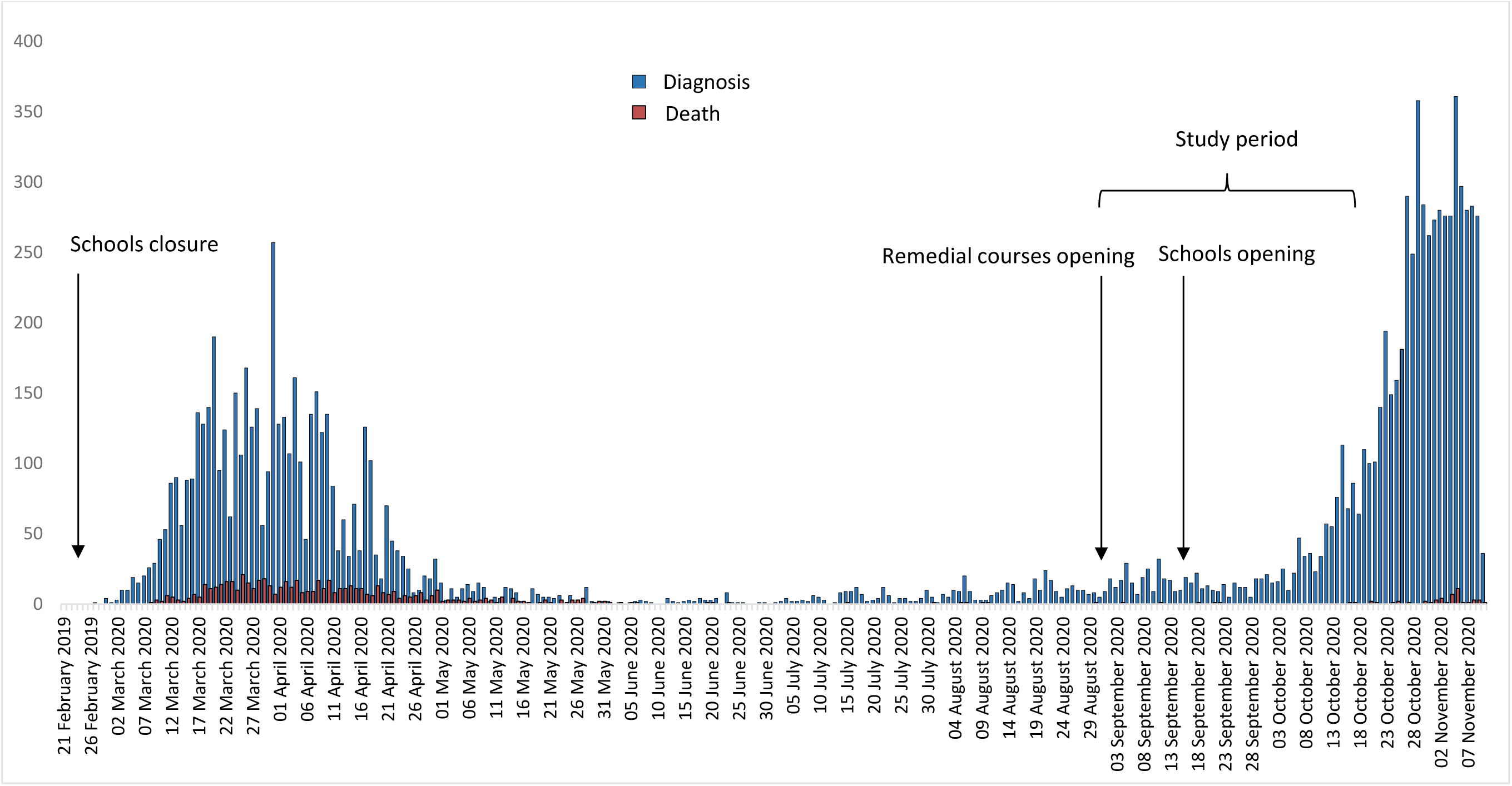
Daily number of notified COVID-19 cases and deaths in Reggio Emilia province from 27 February 2020 (the start of the epidemic in Italy) until 10 November 2020 (11878 cases and 658 deaths).

### Study design

We included in these analyses all consecutive cases leading to a school investigation that were diagnosed between September 1 and October 15 in Reggio Emilia province. We report the characteristics of the school, of the index case, including the possible source of infection, and the number of contacts (students and teachers/personnel) that were identified and tested. When secondary cases were identified, we also provide a description of the cases, with the possible source of infection, to rule out a source of infection outside of school.

The study was approved by the Area Vasta Emilia Nord Ethics Committee (n.2020/0045199).

### Policies of social distancing in schools

During the study period, schools adopted the following social distancing measures: mandatory wearing of surgical masks at all times except when students are seated at their desk and are not speaking; only single desks are used (rather than the traditional double desks), and desks must be at least one meter apart; crowding at separate school entrances and exits is minimized by creating temporal and spatial pathways for the different classes; mixing classes for curricular activities is minimized; extra-curricular activities have all been suspended (12). In some schools, when the classrooms are not big enough to respect social distancing, students are divided into two groups, which alternate attending school and remote learning. Wearing a mask in primary schools is never mandatory (12).

### Contact tracing procedures

All SARS-CoV-2-positive tests performed by the Local Health Authority molecular lab are automatically reported to the Public Health Department, as are the positive lab results of those Reggio Emilia residents who performed the test outside the province. The Local Health Authority starts an epidemiological survey for each case, identifying all close contacts from the 48 hours before symptom onset; these individuals are immediately isolated and tested at least six days after the first contact with the index case, and are retested before the end of the 14 days of quarantine. When a case attends or teaches/personnel at a school, everyone in the class is immediately tested; if the swab is performed earlier than six days from the last contact with the index case, a second swab is also collected at 10/14 days. The epidemiological investigation includes an assessment of the nature of the contact between the index case and his/her classmates, which determines isolation measures: a) all students are isolated if the physical classroom itself makes maintaining distance impossible and/or masks are not worn constantly and/or if secondary cases are ascertained; or b) only those in close contact or who have contact outside of school are isolated, provided that social distancing with the other students has been respected. If classmates are not isolated and didactic activities continue at school (rather than remote), mask wearing is mandatory all the time once a case has been identified.

## Results

### Description of the index cases and their contacts

In the study period, 41 classes in 36 different schools were notified: eight infant-toddler centres and preschools, 10 primary and 18 secondary schools (Table 1). Nine hundred and ninety-four students and 204 teachers were tested during the epidemiological investigations. In all cases, only single classes were considered, except in a preschool, where a larger group (a so-called bubble) sharing common spaces was tested, and in a secondary school, where two classes that did curricular activities together were tested.

**Table 1.** Characteristics of educational/childcare facilities and their students and staff

|  | n | % |
| --- | --- | --- |
| Number of schools | 36 | 100 |
| Type of school |  |  |
| Infant-toddler centre and preschool (0-5 years) | 8 | 22.2 |
| Primary school (6-10 years) | 10 | 27.8 |
| Secondary school | 18 | 50.0 |
| Middle school (11-13 years) | 5 |  |
| High school (14-19 years) | 13 |  |
| Number of classes | 41 |  |
| Index cases | 43 | 100 |
| Students | 38 | 88.4 |
| Teachers | 5 | 11.6 |
| Number of contacts identified during investigations | 1198 | 100 |
| Tested students | 994 | 82.9 |
| Students not tested | 2 | 0.2 |
| Teachers/staff | 204 | 16.8 |
| Secondary cases | 39 | 100 |
| Students | 39 | 100 |
| Teachers/staff | 0 | 0 |

### Secondary attack rate

Thirty-nine secondary cases (3.9%) were identified among 994 children tested, in a total of 13 classes: in one primary school, in five middle schools and in three high schools (Table 2). The attack rate was higher in secondary schools (6.64%) than in primary schools (0.44%), while there were no secondary cases in the preschool settings. There were no secondary cases among tested teachers and staff members.

**Table 2.**
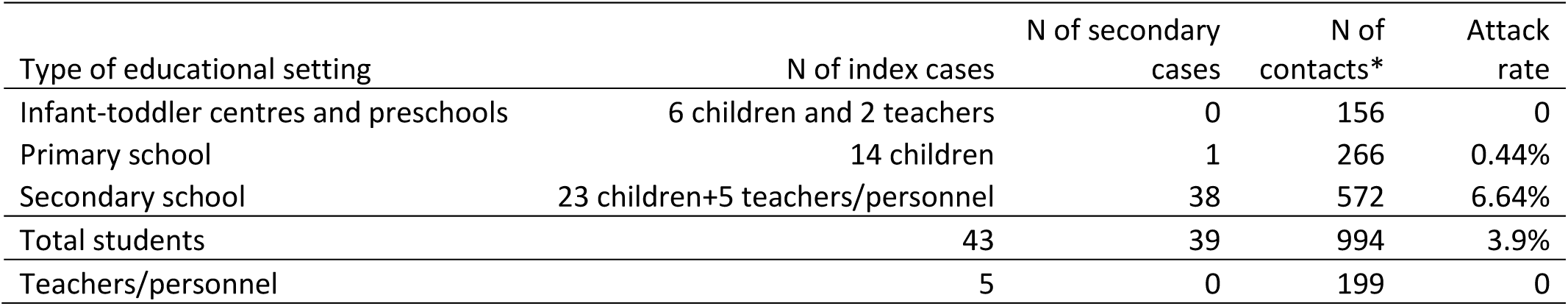
Secondary attack rates by the level of educational facility

### Description of school clusters

Regarding the cluster in the primary school (Cluster 1), the index case had been infected most probably by the relative (Table available upon request-see Data Availability Statement). All the classmates, teachers and school staff were tested; only one asymptomatic case was found.

One of the two clusters (Cluster 2) in a secondary school was identified after a student tested positive; his family member tested positive after returning from a high-incidence area. Both the index case and the family member were asymptomatic. Three positive cases were identified; all developed mild symptoms. No other possible sources of transmission were identified for the three cases.

Investigation of the other cluster in secondary school (Cluster 3) started after the almost simultaneous reporting of two cases in a class: one symptomatic student tested positive and one contact of a family cluster tested positive a day after. All the classmates and teachers were tested and isolated. Six resulted positive, two of whom reporting mild symptoms ten days before positivity of the swab. An analysis of the possible sources outside of school and dates of symptom onset made it possible to identify the asymptomatic case, as the only index case.

Investigations of Clusters 5 and 7 in two different secondary schools started after three siblings (all asymptomatic) reported contact with their symptomatic family member. Both clusters had one secondary case.

In Cluster 6, both the index case and the secondary case had previous contact with a positive person, and the temporal association was difficult to establish.

For Clusters 4 and 8, no possible sources of infection were identified.

The clusters in the middle school (Cluster 9) occurred in several of the schools under the same administration. There were four cases each in two classes and two, five and six in one class each. The index cases were possibly two teachers working at the school. A possible contact outside of school was reported for only one secondary case, but it was not possible to identify the reported case. At least one case in only two classes reported possible sources of infection outside the school, but both classes had also had close contact with the positive teachers. No contacts outside of school were identified for the other three classes, and the dates are compatible with transmission between cases from different classrooms and buildings.

## Discussion

Secondary cases occurred in 13 out of 41 classes, generated from 39 index cases with a secondary attack rate of 3.9%. The largest cluster, with 22 secondary cases, occurred in a middle school. Accordingly, the attack rate was higher in secondary schools than in primary schools, while in the early childhood educational settings as well as among teachers/personnel, secondary transmission was absent.

The inclusion criteria (all consecutive cases), the uniform investigation protocol applied during this phase of the epidemic (testing all classmates) and the population-based nature of the study allowed us to estimate a risk of secondary cases in this context that is not biased by a selection of clusters.

This report is limited by the low number of clusters analysed but has the advantage of an accurate analysis of the chain of transmission, which allowed an assessment of cases with a very high plausibility of transmission occurring from one classmate to another. This made it possible to reasonably rule out other sources. Another limit of these investigations is that they cannot distinguish between transmissions occurring in the classroom and those linked to activities and behaviours outside of school, such as public transports or leisure time. Furthermore, for two cases it was impossible to determine for exactly how many days the students shared the same classroom while the index case was still infectious because he was asymptomatic.

Despite the fact that SARS-CoV-2 incidence was increasing rapidly in this period, this did not interfere with our analyses since we did not identify any other prevalent cases in classes, and all positive cases were most probably secondary cases of the identified index case.

Our data cannot be compared with those recently reported by the Italian COVID-19 surveillance system (6) because our aim was to quantify the risk of secondary transmission in schools and not the incident rate among students and teachers. Contact-tracing studies conducted in schools and educational settings in Australia, Singapore and Ireland found low rates or even no secondary cases (13-15). Only one study reports of screening at the reopening of kindergartens in Korea, which found only one possible secondary case out of 45 cases identified while attending the school (7). Similarly, a low student-to-student attack rate was found in the UK when analyzing predominantly preschools and elementary schools (16) and in Germany for all ages (17). These findings are in line with our report of only one secondary case in elementary schools (out of 10 primary cases) and no secondary case in pre-school (out of 8 primary cases), but not with our results for secondary schools. The policy of not immediately isolating all classmates and delays in testing might explain the difference between our results and those observed in Germany. On the other hand, one large cluster with a high attack rate among students and teachers has been reported in a high school in Israel (18).

Clusters of cases and route of transmission have been investigated in other workplaces whose structural characteristics may be similar to those of schools, i.e. open spaces where people spend several hours every day. Some studies have reported a much higher attack rate of secondary cases than that reported in our series (19, 20). It is worth noting that in most of these cluster analyses, sporadic cases not leading to any known secondary cases are not included, and often only the largest clusters are investigated with well-defined inclusion criteria and scientifically sound methods, allowing publication. Thus, the quality constraints required for scientific publication probably introduced a bias toward overestimating the risk of secondary transmission in workplace settings. Indeed, in a systematic assessment of secondary cases in Brunei, workplaces showed a lower attack rate (<1%) than did households (10.6%) (21).

## Conclusions

Transmission within schools occurred in a non-negligible number of cases, particularly in secondary schools. More timely isolation and testing of classmates and their teachers may likely be effective in reducing virus transmission in this setting.

## Data Availability

According to Italian law, anonymized data can only be made publicly available if there is no potential for the re-identification of individuals (https://www.garanteprivacy.it). Thus, the data underlying this study are available on request to researchers who meet the criteria for access to confidential data. In order to obtain data, approval must be obtained from the Area Vasta Emilia Nord (AVEN) Ethics Committee, who would then authorize us to provide aggregated or anonymized data. Data access requests should be addressed to the Ethics Committee at as well as to the authors at the Epidemiology unit of AUSL - IRCCS of Reggio Emilia at, who are the data guardians.

## Acknowledgements

We thank all the Public Health Department officers and COVID-19 reference persons in the schools that collaborated in the investigations.

We thank Jacqueline M. Costa for the English language editing the text.

## Authors’ contributions

PGR, EL, MV and PP designed the study. EL, MC, SC, EB, FV and EB contributed to the collection and management of the data. OD analysed the data. OD and PGR interpreted the data and wrote the manuscript. All authors revised the manuscript and approved the final version.

